# STREAM: State Trajectory Representation & Evolution-Aware Monitoring

**DOI:** 10.64898/2026.02.03.26345478

**Authors:** Ali Namvar, Sundaresh Ram, Wassim W. Labaki, Stefanie Galban, Njira L. Lugogo, Craig J. Galban

## Abstract

Intensive care unit (ICU) monitoring faces a critical challenge: translating continuous physiological data into actionable insights for real-time clinical decisions. We developed STREAM (State Trajectory Representation & Evolution-Aware Monitoring), which applies optimal transport theory to identify distinct physiological states from routine ICU data and maps individual patients onto state progressions. Using the multicenter eICU Collaborative Research Database (N=158,294), STREAM identified five reproducible states. Patients whose measurements differ substantially from their assigned state (state outliers) showed 9-fold higher mortality (45.7%) versus patients who remained inside state boundaries (5.1%). State-based features predicted mortality with AUROC 0.863 at 8 hours and 0.903 at 72 hours, with excellent calibration error score (0.002). External validation on MIMIC-IV (N=84,517) demonstrated robust performance (AUROC 0.798 and 0.857, respectively), with state outliers exhibited a 7.1-fold higher mortality risk (41.1% vs. 5.8%). Importantly, STREAM connects this risk stratification to the underlying clinical measures defining each physiological state, providing accurate mortality predictions and interpretable insight.

## Introduction

Critical care monitoring faces a fundamental challenge: translating continuous streams of laboratory results, vital signs, and physiological measurements into actionable clinical insights that guide therapeutic decisions. Current ICU monitoring largely, depends on threshold-based alerts for individual parameter abnormalities and composite severity scores (APACHE II, SOFA) that summarize complex physiology into a single daily value (1, 2). While these approaches provide standardized risk assessment, they have critical limitations for real-time clinical decision-making. First, traditional severity scores typically require 24 hours of data collection, delaying risk assessment during the crucial early hours of ICU admission when many therapeutic decisions are made (3). Second, they compress multidimensional physiology into one score, losing information about relationships between organ systems that characterize complex critical illness. Third, they provide static risk estimates without revealing disease trajectory or progression patterns that could guide proactive interventions (4).

Machine learning has produced sophisticated ICU outcome prediction models, with deep learning approaches achieving strong discriminative performance for mortality prediction by processing high-dimensional temporal data (5). However, many of these models function as “black boxes” that output risk probabilities without clinical explanations, limiting their utility for bedside decision-making. Although techniques like SHapley Additive exPlanations (SHAP) provide post-hoc feature importance scores, these model-specific interpretations remain difficult to translate into clear clinical states or actionable physiological insights for individual patients (6–8). Even when such models achieve high predictive performance, clinicians struggle to understand why predictions change over time or which specific interventions would improve patient outcome.

Beyond simply predicting risk (e.g., mortality), clinicians need systems or models that provide a clear structure for understanding how a disease evolves over time, so they can make informed decisions about treatment and intervention. They recognize that critically ill patients follow different pathways through illness: some experience steady improvement, others show gradual deterioration, and some undergo rapid decompensation or unexpected recovery. However, existing monitoring systems lack methods for systematically identifying these progression patterns or predicting when patients might transition between different phases of illness. This gap between clinical intuition about disease progression and available monitoring tools leaves deterioration signals undetected until physiological decompensation becomes clinically apparent, frequently too late for optimal intervention (9, 10).

We developed STREAM (State Trajectory Representation & Evolution-Aware Monitoring), a state-based analysis method that addresses these limitations by applying optimal transport theory (11) to identify distinct multidimensional physiological states from routine ICU data. Unlike approaches that cluster patients once at baseline and then track deviations, STREAM continuously identifies physiological states across time using temporal snapshots of clinical data. A consensus voting algorithm based on optimal transport distances detects meaningful changes in population patterns, indicating state transitions. STREAM’s core innovation lies in mapping individual patients onto these population-derived states to create personalized state progression profiles. This approach enables identification of patients who deviate substantially from typical illness patterns by consistently remaining outside their expected state boundaries. Such state outliers may represent high-risk populations experiencing atypical physiological patterns. This approach preserves multidimensional physiology, discovers states from data without predetermined categories, and explains which variables drive state assignments and transitions.

We evaluate STREAM using the multicenter electronic ICU Collaborative Research Database (eICU-CRD; N=158,294) for development and the single-center Medical Information Mart for Intensive Care IV (MIMIC-IV; N=84,517) database for external validation. STREAM demonstrates strong predictive performance while providing interpretable explanations for each patient’s state progression, identifying high-risk state transitions with clear explanations of the driving physiological factors to support proactive clinical decision-making.

## Results

### Data and Cohort Overview

Figure 1 summarizes the STREAM workflow from raw clinical data to state detection, patient mapping, and downstream applications including early warning signals and outcome prediction. STREAM was developed and evaluated using 158,294 ICU patients from the multicenter eICU-CRD for model development and 84,517 patients from the single-center MIMIC-IV database for external validation (Supplementary Table 1). Both cohorts had similar demographic characteristics and severity of illness distributions, supporting the validity of cross-dataset comparison.

**Figure 1.**
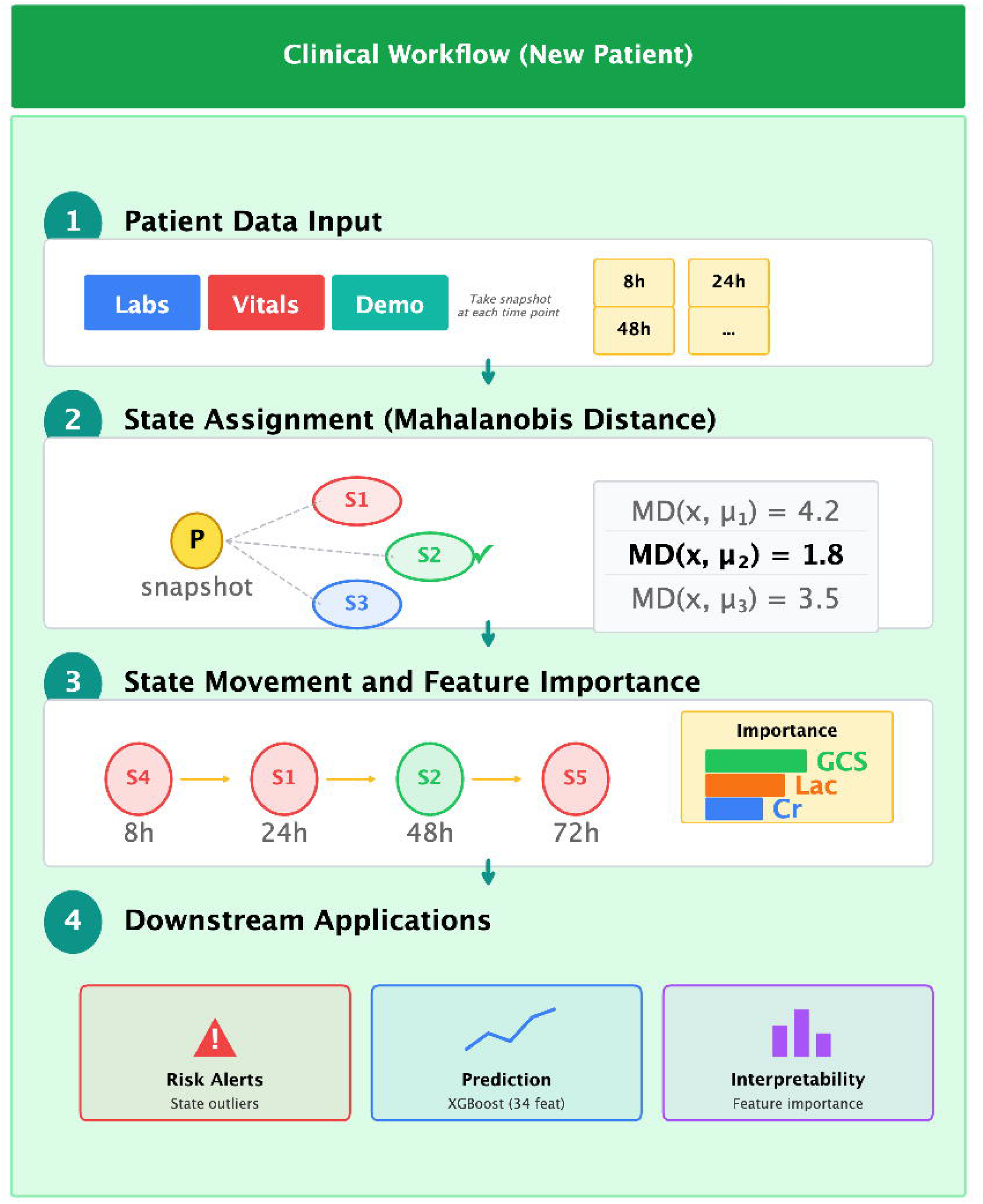
STREAM Clinical Workflow. Schematic representation of the STREAM framework for processing new patient data. (1) Patient Data Input: laboratory values (Labs), vital signs (Vitals), and demographics (Demo) are collected and processed at each time window (8h, 24h, 48h, etc.). (2) State Assignment: each patient snapshot is assigned to one of five population states (S1-S5) using Mahalanobis distance (MD) to state centroids; the state with minimum MD is selected (example shows S2 selected with MD = 1.8). (3) State Movement and Feature Importance: At each snapshot, the patient’s state assignment (example: S4 → S1 → S2 → S5) and feature importance (e.g., GCS, lactate, creatinine at 48hrs) are determined. (4) Downstream Applications: STREAM outputs support three clinical functions: risk alerts based on state outlier detection, mortality prediction via XGBoost models using 34 features, and interpretability through feature importance analysis. Abbreviations: MD, Mahalanobis distance; S1-S5, physiological states 1-5; GCS, Glasgow Coma Scale; Lac, lactate; Cr, creatinine; Demo, demographics.

Twenty-eight routinely collected measurements spanning organ function (kidney, liver, hematologic), metabolic status (glucose, electrolytes, acid–base balance), and monitored physiology (hemodynamics, respiratory function, neurologic status) were used to develop STREAM (Table 1). Each ICU stay was represented by time-based snapshots covering a pre-admission window, followed by snapshots at 8, 16, and 24 hours, then at 48 and 72 hours, and daily from day 4 to day 10, yielding longitudinal profiles of each patient’s clinical data. These longitudinal profiles formed the foundation for state discovery and individualized patient mapping reported below.

**Table 1.**
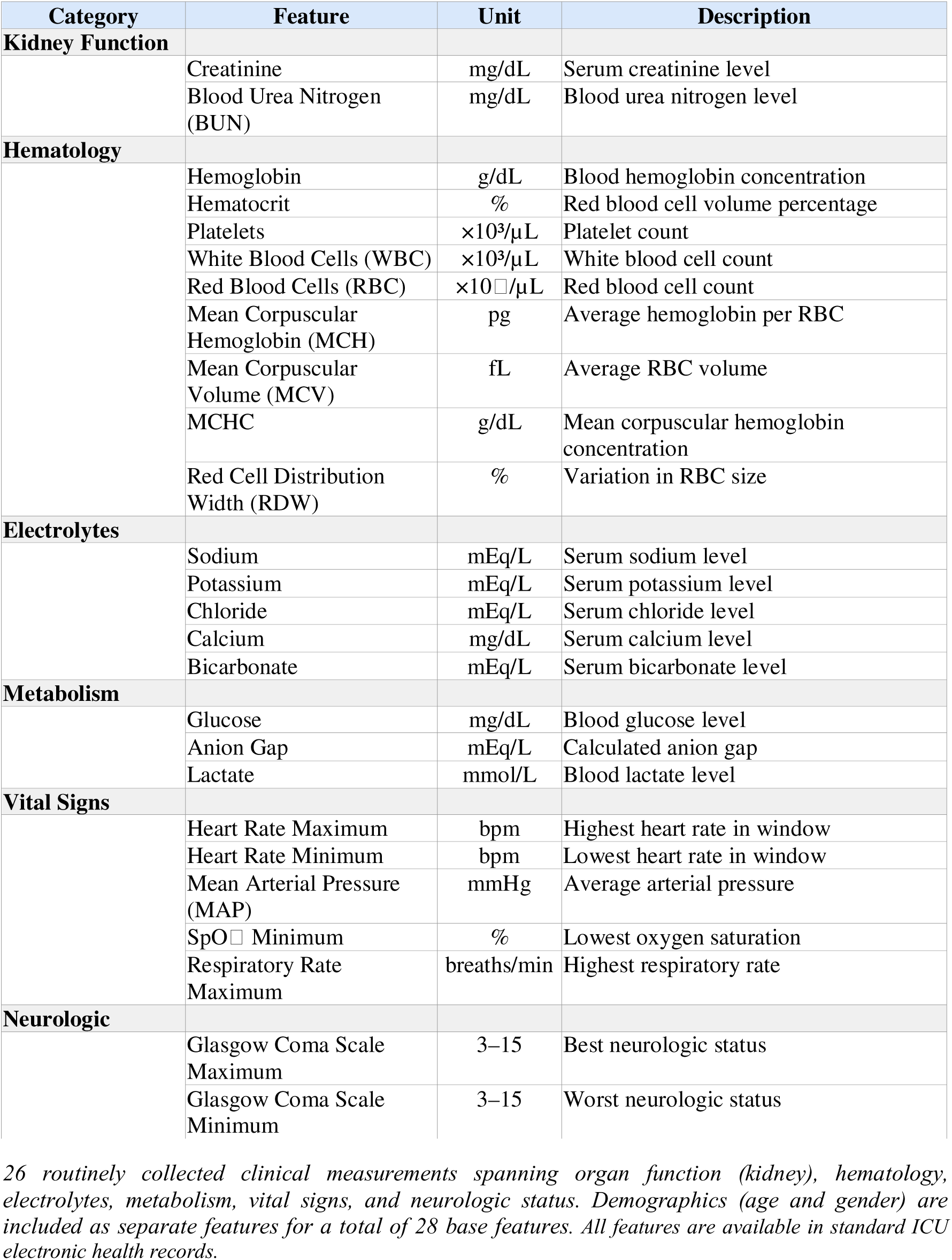
Clinical features used in STREAM.

### STREAM Physiological State Discovery

Five distinct physiological states were identified across the ICU course in eICU-CRD (Fig. 2A) using a voting system that combines three distance metrics from optimal transport theory. These states represent physiological patterns detected at different snapshots and are not hierarchical in nature. The first transition between states was detected within 24 hours of ICU admission, with subsequent transitions occurring at day 3 (72 hours), 6 and 8 of the ICU stay.

**Figure 2.**
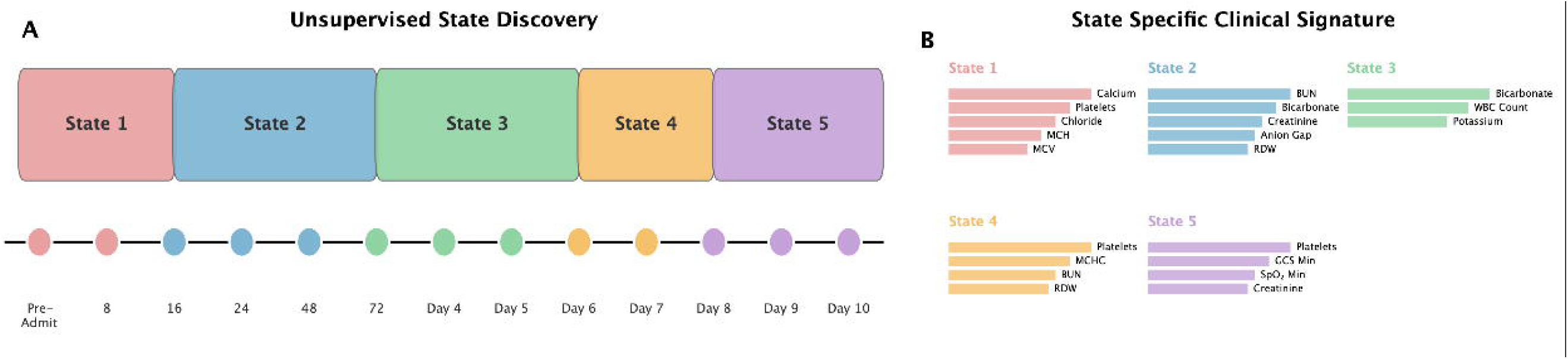
Physiological State Discovery and Characterization in eICU-CRD. **(A)** Unsupervised state discovery. STREAM identifies five recurrent physiological states (States 1–5) across ICU snapshots from pre-admission through day 10. **(B)** State clinical signatures. Bar plots show the top variables characterizing each state. State 1: calcium, platelets, chloride, MCH, MCV. State 2: BUN, bicarbonate, creatinine, anion gap, RDW. State 3: bicarbonate, WBC, potassium. State 4: platelets, MCHC, BUN, RDW. State 5: platelets, GCS minimum, SpO□ minimum, creatinine. *Abbreviations: BUN, blood urea nitrogen; WBC, white blood cell count; MCHC, mean corpuscular hemoglobin concentration; GCS, Glasgow Coma Scale; SpO*□*, oxygen saturation; MCH, mean corpuscular hemoglobin; MCV, mean corpuscular volume; RDW, red cell distribution width*.

Each state exhibited a distinct clinical signature reflecting characteristic patterns of critical illness progression (Fig. 2B). State 1 was characterized by electrolyte and hematologic abnormalities including calcium, chloride disturbances, and red cell indices (MCH, MCV, platelets).(12). State 2 showed elevations in renal and metabolic markers including BUN, creatinine, bicarbonate, anion gap, and RDW. State 3 was characterized by inflammatory and metabolic features including elevated WBC count, bicarbonate, and potassium abnormalities(13). State 4 demonstrated hematologic changes with platelets, MCHC, BUN, and RDW elevations. State 5 showed multi-organ involvement with depressed Glasgow Coma Scale scores, low SpO□, elevated creatinine, and platelet abnormalities (14, 15).

### Patient State Mapping, Alignment, and Feature Importance

Throughout the ICU stay, patients were assigned to the nearest state based on Mahalanobis distance from state centroids at every snapshot, creating a state progression pathway. For each state, we identified all unique patients who entered that state at any time during their ICU course and calculated the proportion who died in ICU. State 5 had the highest mortality rate (10.1%), followed by State 1 (9.0%) (Fig. 3A). States 2, 4, and 3 had lower mortality rates (4.5%, 4.1%, and 6.6%, respectively). Notably, although States 1 and 5 showed similar mortality rates, their clinical signatures differ markedly (Fig. 2B): State 1 is characterized by hematologic and electrolyte abnormalities, whereas State 5 reflects multi-organ failure with neurologic and respiratory compromise.

**Figure 3.**
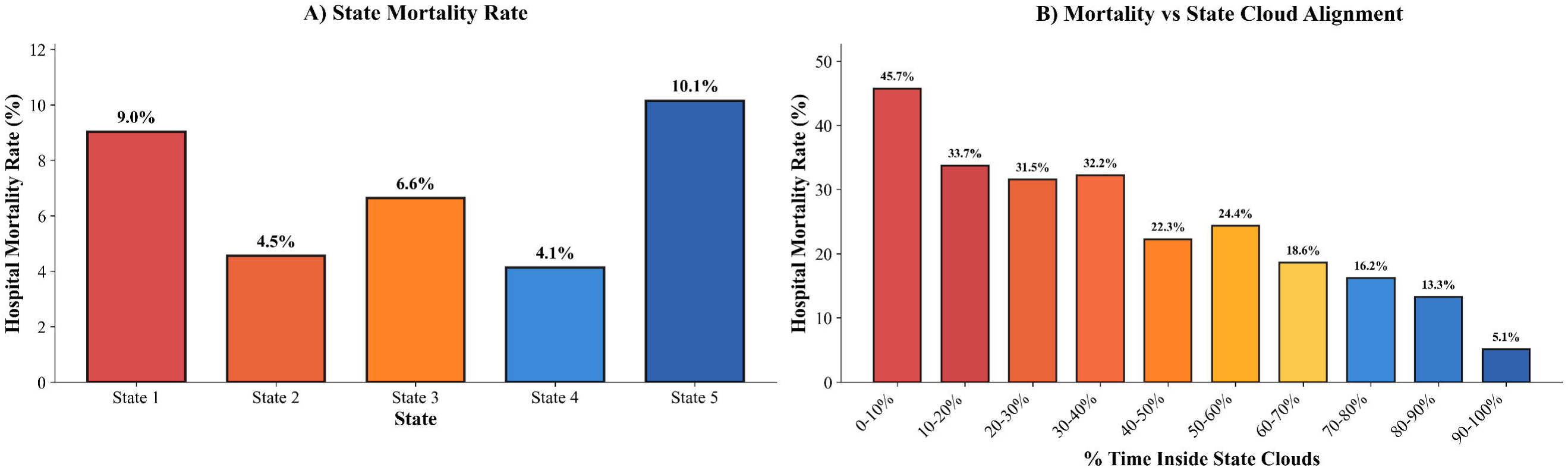
State Mortality and Geometric Alignment Patterns in eICU-CRD. **(A)** Hospital mortality by states visited during the ICU course. State 5 (10.1%), State 1 (9.0%), State 3 (6.6%), State 2 (4.5%), State 4 (4.1%). **(B)** Mortality versus state alignment. Mortality increases as alignment to assigned state clouds decreases (percent of ICU time inside the 95% state boundaries). State outliers (0–10% alignment) have 45.7% mortality versus 5.1% for patients with 90–100% alignment (9-fold difference). State-specific outlier rates are in Supplementary Results S.R.4.

in addition to patient State assignment, we determined how well each patient’s measurements matched their assigned state. We defined state outliers as patients who spent minimal time within the 95% confidence boundary of their assigned state, indicating physiological patterns that deviated substantially from typical state characteristics. Mortality was 45.7% in patients with 0-10% alignment compared with 5.1% for patients who remained inside state boundaries (90-100% alignment). Mortality decreased progressively as alignment increased (Fig. 3B). Notably, state outliers showed elevated mortality across all five states, regardless of whether the state itself was low-risk or high-risk (Supplementary Results S.R.4).

Figure 4 illustrates a representative patient that demonstrates a high-risk state progression with feature importance profiles. At ICU admission, the patient was assigned to State 1 as a state outlier, remaining outside state boundaries during the first 24 hours with high Mahalanobis distance with early drivers included metabolic stress (potassium and bicarbonate abnormalities) (Fig. 4A–B). By day 3, the patient transitioned to State 5, where hematologic changes predominated (falling hemoglobin and platelet counts). Although the patient was inside the state boundaries on days 4 and 5, conditions returned to State 5 (days 6–8), where the patient remained until death. Feature importance profiles reveal the clinical drivers at each snapshot. Notably, feature importance changed even when the patient remained in the same state. While in State 1, for example, dominant features shifted from potassium and RDW at admission to glucose and anion gap by day 3, demonstrating that STREAM can detect evolving pathophysiology before state transitions occur.

**Figure 4.**
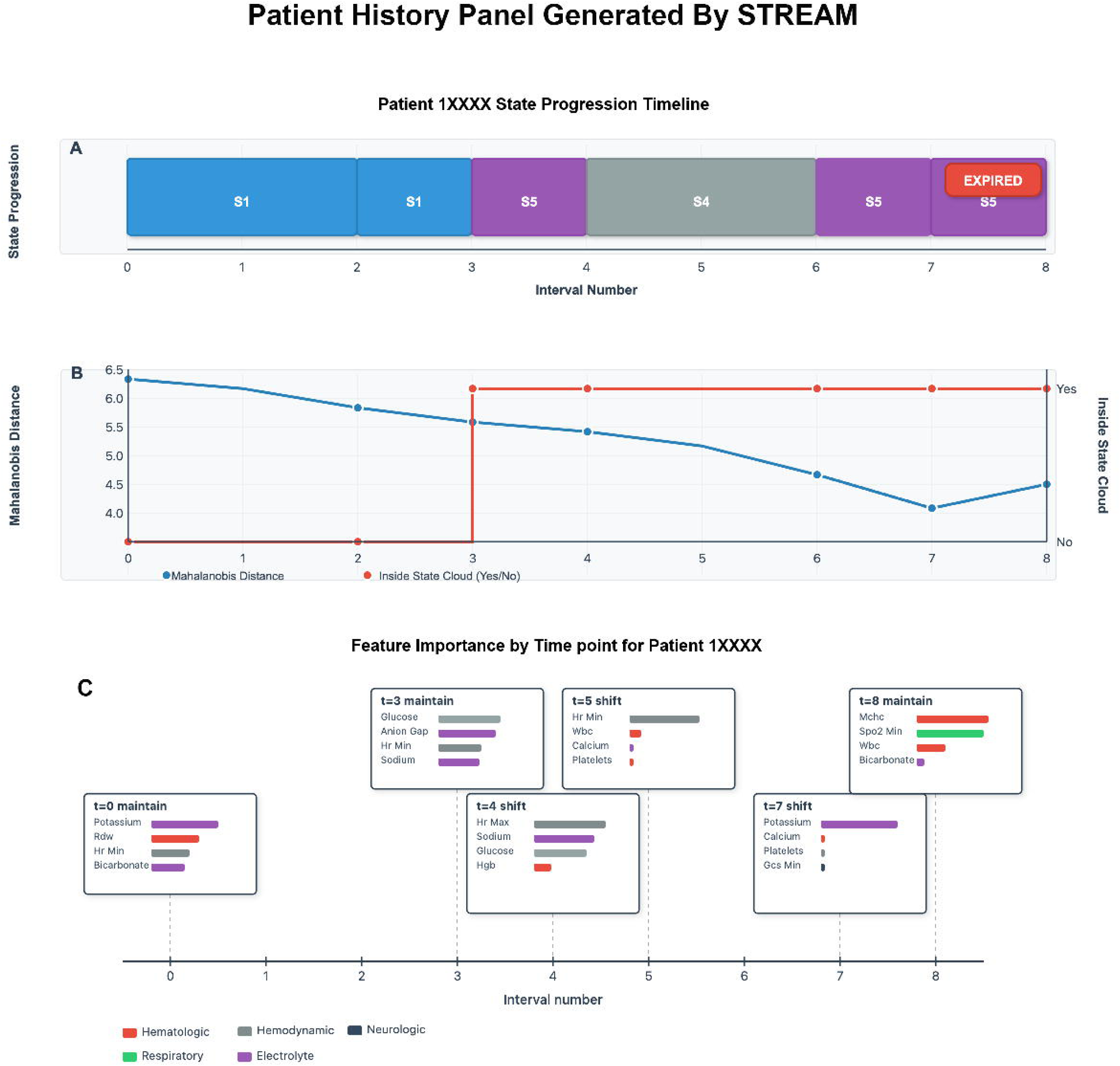
Patient History Panel Generated by STREAM. Example high-risk patient state movement. **(A)** State progression across ICU intervals (State 1 → State 5 → State 4 → State 5) with death. **(B)** Geometric alignment over time. Mahalanobis distance to the assigned state centroid with in-cloud status (inside/outside the 95% state boundary). **(C)** Time-resolved feature importance highlighting variables driving state maintenance or shifts at selected intervals. Feature colors denote physiological domains (hematologic, hemodynamic, neurologic, respiratory, electrolyte).

### Mortality Prediction Using Gradient Boost Model

XGBoost (extreme gradient boosting) models trained with STREAM-derived state geometry features predicted ICU mortality in eICU-CRD (Supplementary Table 2) cohort. To ensure that predicted probabilities reflected observed outcome frequencies, we applied post-hoc calibration to our model using Platt scaling and isotonic regression. Calibration substantially improved, with expected calibration error decreasing from 0.037 (uncalibrated) to 0.005 with Platt scaling and to 0.002 with isotonic regression (Supplementary Results S.R.1; Supplementary Fig. 1). Discriminative performance was strong across time horizons: area under the receiver operating characteristic curves (AUROC) was 0.863 at 8 hours, 0.873 at 24 hours, 0.890 at 48 hours, 0.903 at 72 hours, and 0.932 using complete ICU stay data (Table 2). Corresponding area under the precision-recall curve (AUPRC) ranged from 0.368 at 8 hours to 0.561 for full stay.

**Table 2.** Predictive performance of STREAM-based mortality models in eICU-CRD.

| Time Window | Outcome | AUROC (95% CI) | AUPRC (95% CI) |
| --- | --- | --- | --- |
| 8 hours | ICU Mortality | 0.863 (0.860–0.866) | 0.368 (0.360–0.375) |
| 24 hours |  | 0.873 (0.868–0.878) | 0.401 (0.386–0.416) |
| 48 hours |  | 0.890 (0.886–0.893) | 0.452 (0.436–0.468) |
| 72 hours |  | 0.903 (0.898–0.908) | 0.485 (0.474–0.495) |
| Full Stay |  | 0.932 (0.930–0.933) | 0.561 (0.549–0.573) |

*Performance of XGBoost models trained on STREAM-derived features for predicting ICU mortality. Each model used patient-level cross-validation with early stopping. AUROC, area under the receiver operating characteristic curve; AUPRC, area under the precision-recall curve.*

We performed a SHapley Additive exPlanations (SHAP), analysis to identify the features that most strongly influenced mortality predictions (Fig. 5; Supplementary Results S.R.2; Supplementary Fig. 2). At 8 hours, clinical variables such as lactate dominated predictions (Fig. 5A). By 72 hours, state geometry features, particularly Mahalanobis distance, emerged as the strongest contributor (Fig. 5B), demonstrating the increasing value of longitudinal, trajectory-based information as more temporal data accrued. Across all evaluated time windows, state geometry features, including mean, minimum, and maximum Mahalanobis distance, consistently ranked among the top predictors and frequently surpassed the importance of individual physiological measurements. Age and Glasgow Coma Scale values also remained influential, alongside markers of renal function (creatinine, BUN), inflammatory markers (WBC), and metabolic stress (lactate).

**Figure 5.**
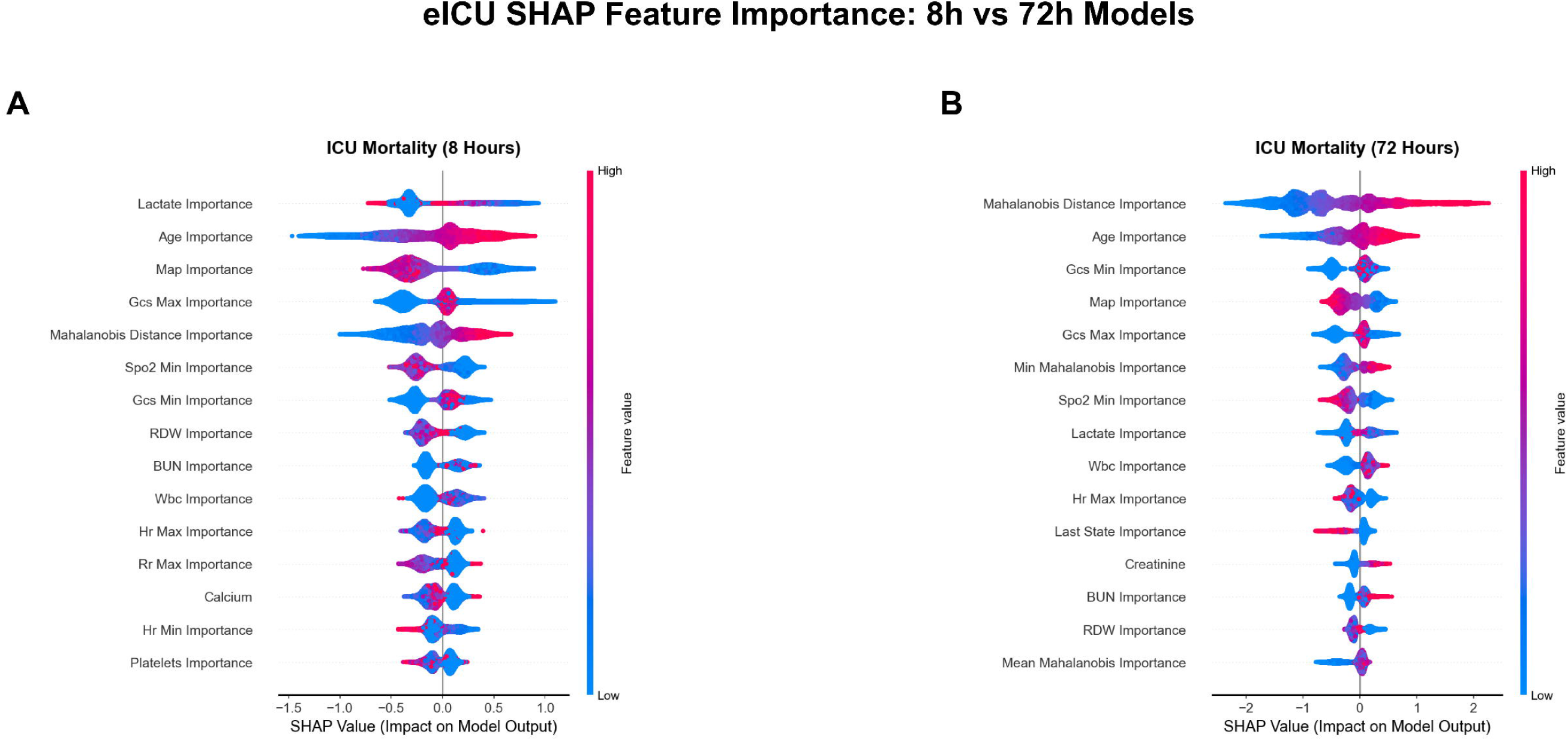
SHAP Feature Importance Analysis Comparing Early and Extended Prediction Horizons. **(A)** 8-hour mortality model. Top features by mean |SHAP| include lactate, age, MAP, GCS maximum, Mahalanobis distance, and SpO□ minimum. **(B)** 72-hour mortality model. Mahalanobis distance becomes the top feature, with age, GCS, MAP, and SpO□ remaining prominent. SHAP summary plots show per-patient contributions (x-axis) for each feature (rows), with color indicating feature value.

For completeness, we compared STREAM prediction model with a baseline model using only raw clinical features. Models that employ STREAM defined state geometry features consistently outperformed baseline models across all time horizons (Supplementary Table 3; Supplementary Results S.R.6).

### External Validation in MIMIC-IV

To assess generalizability, we applied the eICU-trained model to the independent MIMIC-IV dataset (N=84,517). Cross-dataset application required quantile-to-quantile distribution alignment to address systematic measurement differences between datasets (Supplementary Results S.R.3; Supplementary Table 4).

State-specific mortality patterns replicated in MIMIC-IV (Fig. 6A): State 5 showed highest mortality (11.6% vs 10.1% in eICU-CRD), followed by State 1 (10.5% vs 9.0%), while States 2 and 4 remained lowest-risk. The state outlier-mortality relationship also replicated (Fig. 6B): state outliers (0-10% time inside state boundaries) had 41.1% mortality versus 5.8% for patients who remained inside state boundaries (90-100%), a 7.1-fold difference. State outlier mortality rates were consistent across all states, confirming that state outlier status confers elevated risk independent of state assignment (Supplementary Results S.R.4).

**Figure 6.**
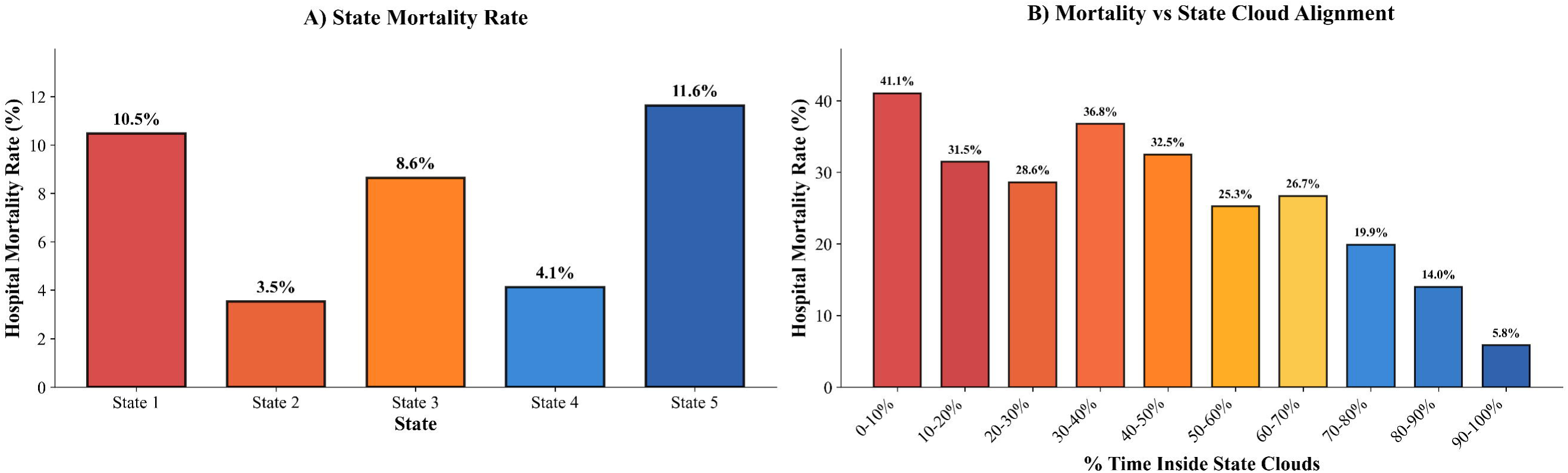
Cross-Dataset Validation of State Mortality and Geometric Alignment Patterns in MIMIC-IV. **(A)** Mortality by states visited when MIMIC-IV patients are projected onto eICU-derived state boundaries. State 5 (11.6%), State 1 (10.5%), State 3 (8.6%), State 4 (4.1%), State 2 (3.5%). **(B)** Mortality versus state alignment. Outliers (0–10% alignment) have 41.1% mortality versus 5.8% for 90–100% alignment (7.1-fold difference), reproducing the alignment–mortality gradient. State-specific outlier rates are in Supplementary Results S.R.4.

The eICU-trained model maintained strong performance without retraining (Table 3). AUROC decreased by only 3–7% across all time windows: 0.798 at 8 hours, 0.815 at 24 hours, 0.836 at 48 hours, 0.857 at 72 hours, and 0.899 for full stay (Supplementary Results S.R.4). Geometric alignment features remained among top predictors in both datasets (Supplementary Results S.R.5; Supplementary Fig. 3).

**Table 3.** Generalization to MIMIC-IV: performance of STREAM-based mortality prediction.

| Outcome | Time Window | MIMIC-IV AUROC<br>(95% CI) | eICU-CRD<br>AUROC | $\Delta$ AUROC |
| --- | --- | --- | --- | --- |
| ICU Mortality | 8 hours | 0.798 (0.796–0.799) | 0.863 | −0.065 |
|  | 24 hours | 0.815 (0.813–0.816) | 0.873 | −0.058 |
|  | 48 hours | 0.836 (0.835–0.838) | 0.890 | −0.054 |
|  | 72 hours | 0.857 (0.857–0.858) | 0.903 | −0.046 |
|  | Full Stay | 0.899 (0.899–0.900) | 0.932 | −0.033 |

*Performance of eICU-trained XGBoost models applied to MIMIC-IV without retraining. $\Delta$ AUROC indicates the difference between MIMIC-IV and eICU-CRD performance.*

## Discussion

Critical care monitoring currently relies on threshold-based alerts and composite scores that compress multidimensional physiology into a single value, limiting their ability to capture evolving patterns of organ dysfunction. STREAM offers an alternative paradigm by using optimal transport theory to define data-driven physiological states and track how individual patients move through this state space over time using routinely collected ICU variables. STREAM first assigns each patient snapshot to the nearest physiological state using Mahalanobis distance and then uses this geometry to identify state outliers, patients whose measurements deviate substantially from the characteristic pattern of their assigned state. With strong predictive performance, calibration, and external validation, these findings support geometric, state-based monitoring as a promising framework for bedside decision support.

A unique feature of STREAM is its use of optimal transport theory to stratify clinically relevant states over time. While optimal transport has been applied in medical imaging (for example, brain morphometry classification) and computational biology (for example, single-cell trajectory inference), its use in critical care remains limited. To our knowledge, only Wang and colleagues have applied optimal transport in this setting, using Sinkhorn distance for cross-hospital sepsis domain adaptation, a fundamentally different application than physiological state identification(16). As shown in Figure 2, five distinct physiological states emerged from unsupervised analysis of ICU data, each with distinct clinical signatures and mortality risks ranging from 4.1% in State 4 to 10.1% in State 5. States with similar mortality can still differ clinically; for example, States 2 and 4 have comparable mortality (4.5% versus 4.1%) but distinct signatures, with State 2 marked by elevations in BUN, bicarbonate, creatinine, anion gap, and RDW, and State 4 characterized by abnormalities in platelets, MCHC, BUN, and RDW. STREAM provides patient-specific feature importance that identifies which biomarkers drive each individual’s state assignment and transitions. For state transitions, STREAM quantifies which features caused the change; for patients remaining in the same state, it tracks whether the feature importance profile is stable or shifting over time. This distinction is clinically meaningful: two patients in State 1 may have different profiles (for example, one dominated by thrombocytopenia and hypocalcemia, another by macrocytosis and hyperchloremia), and a single patient’s profile can change substantially while state classification remains unchanged, as illustrated by the evolving feature importance pattern in Figure 4C despite periods of state maintenance.

In addition to state assignment, STREAM also designates individual cases as state outliers. These are patients whose measurements fall outside the 95% confidence boundary of their assigned state. State outliers experienced higher mortality (45.7%) compared with 5.1% for patients who remained inside state boundaries, representing a 9-fold risk difference. This represents a large separation in mortality risk. Prior phenotyping studies have reported smaller separations in other settings. However, these are not direct comparisons, because cohorts, endpoints, and study designs differ. In ARDS, Calfee et al. identified inflammatory sub-phenotypes with about a 2-fold difference in 90-day mortality, and treatment effects differed by subtype. In sepsis, temperature-trajectory sub-phenotypes showed 4-fold higher odds of mortality for the hypothermic group versus the reference group (17, 18). Prior work using routine laboratory panels shows that mortality risk is carried by the overall pattern across tests, not by any single marker alone. Lind and colleagues showed that adding a set of common clinical chemistry tests to standard risk factors improved mortality prediction in two large cohorts (19). Our findings extend this idea to the STREAM state space. A patient can become a state outlier without one lab being severely abnormal. Several mildly abnormal values can add up and shift the patient far from the state center, capturing risk that single threshold alerts may miss. State outlier status appears to be a marker of clinical instability that replicated across both datasets (Supplementary Results S.R.4).

STREAM’s predictive performance compares well with contemporary machine learning early warning approaches when viewed in the context of published results (Table 4). At 8 hours, STREAM achieved AUROC 0.863 (95% CI 0.860 to 0.866), providing actionable risk stratification before traditional severity scores can be calculated. Reported AUROCs in related studies include PICTURE (AUROC 0.819)(20), eCART (AUROC 0.801) (21), HAVEN (AUROC 0.901) (22, 23), and iMORS (internal AUROC 0.964, external 0.870–0.890) (24–26). STREAM showed a modest reduction on external validation. This magnitude of drop is smaller than reductions reported in other cross cohort settings, including an 18% AUROC reduction reported by Chen et al. For sepsis associated acute kidney injury prediction and a 7 to 10% reduction reported for iMORS across external cohorts. STREAM decreased by only 3 to 7% between eICU-CRD and MIMIC-IV. These values are not direct comparisons, because the studies use different endpoints, prediction windows, and cohorts. Taken together, this level of detail may support several clinical applications, including tracking drug response, identifying which organ system to target for treatment, and providing clear explanations for discussions with patients and families about goals of care.

**Table 4.** Comparison of STREAM with existing ICU risk prediction models.

| Model | Year | Internal AUROC | External AUROC | $\Delta$ AUROC | Key Feature |
| --- | --- | --- | --- | --- | --- |
| STREAM (72 h) | 2025 | 0.903 | 0.857 | ~3.5% | State Based OT |
| STREAM (8 hours) | 2025 | 0.863 | 0.798 | ~7.5% | Early prediction |
| iMORS | 2024 | 0.964 | 0.870–0.890 | ~7–10% | Ensemble DL+LGBM |
| XMI-ICU | 2025 | 0.910 | 0.85–0.88 | ~5% | MI patients only |
| eCART | 2014 | 0.801 | 0.75–0.80 | ~5% | Ward patients |
| PICTURE | 2021 | 0.819 | — | — | XGBoost+SHAP |
| HAVEN | 2021 | 0.901 | ~0.88 | ~3% | UK hospitals |
| Epic EDI | 2020 | 0.65–0.80 | Variable | Variable | Proprietary |
| APACHE II | 1985 | 0.58–0.74 | 0.58–0.74 | — | 14 variables |
| APACHE IV | 2006 | 0.76–0.88 | 0.76–0.88 | — | 142 variables |
| SOFA | 1996 | 0.69–0.82 | 0.69–0.82 | — | 6 organ systems |

Abbreviations: OT, optimal transport; DL, deep learning; LGBM, LightGBM; MI, myocardial infarction; EDI, Epic Deterioration Index. Values for comparator models extracted from original publications. STREAM rows highlighted in green.

We acknowledge limitations to this study. We used only retrospective data from US databases, so generalization to international populations and prospective clinical settings will require further evaluation, especially because temporal, geographical, and domain generalizability represent distinct validation requirements. In addition, the current framework analyzes only routinely collected structured clinical data and does not incorporate richer sources such as ventilator waveforms, medical imaging, or unstructured clinical documentation, including physician progress notes and nursing documentation, that might further refine state definitions.

STREAM establishes state-based geometric analysis as a practical framework for ICU monitoring. By combining optimal transport-derived states, patient-level mapping, feature importance, and state outlier detection, STREAM links risk stratification to the clinical measurements that define each physiological pattern. Strong and well-calibrated predictive performance, limited loss of accuracy on external validation, and a consistent association between state alignment and mortality support the potential of state-aware, geometry-based monitoring to complement static severity scores and single-parameter alerts. Because STREAM relies exclusively on routinely collected longitudinal clinical data, the framework is, in principle, extensible to settings such as emergency departments, general wards, chronic disease management, and post-surgical recovery. However, state geometries would need to be learned and validated separately for each new clinical context. Future prospective studies should test whether integrating STREAM into real-time workflows, particularly by identifying state outliers and their driving features, can support earlier and more targeted clinical attention and may improve care efficiency in critical care.

## Methods

### Data Sources and Study Population

We used eICU-CRD version 2.0 for model development and internal validation. This multicenter database contains 200,859 ICU admissions from 335 ICUs across 208 US hospitals between 2014 and 2015, representing diverse practice settings including academic medical centers, community hospitals, and specialty facilities spanning medical, surgical, cardiac, and neurological intensive care units. The database includes minute-by-minute vital signs, laboratory results, medications, and clinical notes, with a median ICU length of stay of 1.9 days (IQR: 1.0-3.8 days). We included adult patients (≥18 years) with ICU stays exceeding 8 hours, yielding 158,294 ICU admissions for analysis. We selected this dataset to demonstrate STREAM’s capability to identify physiological states and predict outcomes using only routine clinical measurements available in standard ICU care, without requiring specialized monitoring or research-grade data collection. MIMIC-IV was used exclusively for external validation. This single-center database contains 76,540 ICU admissions from Beth Israel Deaconess Medical Center (Boston, MA) between 2008 and 2019, representing a large academic medical center with medical, surgical, trauma, and cardiac ICUs. After applying identical inclusion criteria (age ≥18 years, ICU stay >8 hours), we analyzed 84,517 ICU patients. MIMIC-IV provides an independent test of validation across different hospitals, geographic regions, patient populations, and time periods (27, 28).

### Feature Selection and Temporal Structure

#### Clinical measurements

We analyzed 28 routinely collected measurements to represent critical organ systems without requiring specialized equipment (Table 1). To identify the most frequently measured parameters, we ranked all laboratory tests by measurement frequency across the eICU-CRD cohort. We selected the top 19 most frequently measured laboratory values, then added routinely monitored vital signs and neurologic assessments including heart rate extrema (maximum and minimum), mean arterial pressure, oxygen saturation minimum, respiratory rate maximum, and Glasgow Coma Scale extrema (maximum and minimum). This yielded 26 features spanning kidney function (creatinine, blood urea nitrogen), hematology (hemoglobin, hematocrit, platelets, white blood cells, red blood cells, red cell indices), electrolytes (sodium, potassium, chloride, calcium, bicarbonate), metabolism (glucose, anion gap, lactate), and cardiorespiratory-neurologic status. Among demographic data, age and gender were selected. We excluded composite severity scores (APACHE, SOFA) and outcome indicators to prevent the model from learning direct associations with labels rather than underlying physiological patterns.

#### Snapshot creation

Each patient’s timeline began at ICU admission (time zero) and was expressed in hours. We divided each ICU stay into time-based snapshots: one pre-admission snapshot (−24 to 0 hours, capturing emergency department or ward data, if available), snapshots at 8, 16, and 24 hours, followed by snapshots at 48 and 72 hours, and daily snapshots from day 4 to day 10. This structure provides fine temporal resolution during the critical first day when many clinical decisions occur, followed by daily resolution to track longer-term trajectories. Clinical measurements were assigned to snapshots based on timestamp, and all values within each snapshot were averaged per feature to create one physiologic profile per snapshot.

### Data Preprocessing

#### Missing data imputation

ICU data contain substantial missingness due to selective laboratory ordering based on clinical need. We used Multiple Imputation by Chained Equations (MICE) to estimate missing values while preserving relationships between variables (29). For each feature type, we applied appropriate imputation methods: continuous laboratory values used predictive mean matching, categorical variables used mode-based selection, ordinal variables used proportional odds models, and binary indicators used logistic regression. For time-varying measurements such as Glasgow Coma Scale and vital signs, interior gaps were filled by linear interpolation, and the most recent value was extended forward or the earliest value backward to maintain clinical continuity (Supplementary Methods S.M.1).

#### Feature standardization

After imputation, we standardized all features to comparable scales. Continuous laboratory values and vital signs were scaled to [0,1] using minimum and maximum values observed in the eICU-CRD cohort. Ordinal features (e.g., categorical laboratory interpretations) were mapped to [0,1] according to their natural ordering. Binary features remained as 0 or 1. All standardization parameters (minimum, maximum, means) were calculated exclusively from eICU-CRD training data (30).

### Distance-Based State Detection

#### Overview

STREAM detects when the population transitions between physiological states by monitoring changes at three different levels: individual laboratory values, overall patient condition, and patient similarity patterns. We call this the 3-OT (Triple Optimal Transport) system, which operates like three independent judges examining the same data from different perspectives.

#### The Three Detection Methods

OT-1D uses 1-Wasserstein distance computed separately for each of the 28 clinical features between consecutive snapshots (31).This univariate optimal transport metric quantifies distributional shifts in individual biomarkers across the population, detecting feature-specific transitions such as systematic increases in lactate distributions or widening of mean arterial pressure distributions. OT-2D uses the 2-Wasserstein distance to measure movement in the complete 28-dimensional physiological space between consecutive snapshots (32). This multivariate optimal transport metric quantifies the minimum work required to transform one snapshot’s patient distribution into the next, capturing coordinated changes across multiple organ systems simultaneously. OT-Geometric uses Gromov-Wasserstein distance to measure changes in the internal geometric structure of patient populations across consecutive snapshots (33).

#### Consensus Voting System

Each method generates a distance between consecutive snapshots. To determine which time points represent true state transitions rather than random fluctuations, each method independently applies three unsupervised threshold-detection algorithms: KneeLocator (elbow detection) (34), Gaussian Mixture Models (cluster intersection) (35), and K-means clustering (midpoint boundaries) (36). Each algorithm produces a binary (yes/no) vote for each snapshot. The three algorithm votes are aggregated to produce one single vote per detection method. Final transition decisions are made by combining the three OT method votes using confidence-weighted consensus score, defined as:

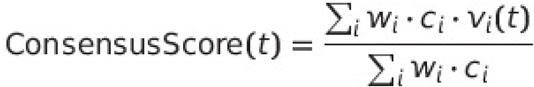

where *V_i_*(*t*)∈{0,1} is the binary vote from method *i* at snapshot *t*, *W_i_*is the base weight, and *C_i_* is a confidence score calculated from each method’s signal dynamic range and consistency across snapshots. A state transition is recorded when ConsensusScore(*t*) > 0.5, corresponding to agreement from at least two of the three methods.

#### Implementation for eICU-CRD

For the multicenter eICU-CRD cohort, we used a consensus threshold of 0.5, requiring agreement from at least two of the three methods. This choice accounts for heterogeneity across hospitals and patient populations while balancing sensitivity to avoid false positives driven by isolated fluctuations in individual methods.

### Individual Patient Mapping and State Assignment

After identifying population-level states using the 3-OT voting system, each patient snapshot was assigned to its nearest state to track individual state progressions through the ICU course. For each detected state, a centroid (mean feature vector) was calculated in the 28-dimensional feature space, and an inverse covariance matrix was computed to capture the state’s characteristic variance structure.

Patient assignment used Mahalanobis distance to account for feature correlations and variable scales within each state (37). For each patient at each snapshot, Mahalanobis distance was calculated from the patient’s feature vector to each state centroid. Patients were assigned to the state with the smallest Mahalanobis distance, generating a sequence of state labels over time that constitutes the patient’s state progression pathway.

Patient-level alignment metrics were computed to quantify how well each patient matched their assigned state. At each snapshot, we calculated the normalized Mahalanobis distance and determined whether the patient fell inside or outside the 95% confidence boundary of their assigned state. Alignment was defined as the percentage of snapshots in which a patient remained inside the boundaries of their assigned states across their ICU stay. State outliers were defined as patients with low alignment (≤10% of time inside state boundaries), indicating substantial deviation from typical state patterns.

### Feature Importance Calculation

Following state assignment at each snapshot, we computed snapshot-specific feature importance scores to quantify each clinical variable’s contribution to individual patient state progressions. We developed a hybrid importance scoring system that adapts to different state progression patterns while maintaining interpretability (38).

The system distinguishes between two clinical scenarios. For patients who remained in the same state across consecutive snapshots, maintenance importance quantifies which features characterize a stable physiological pattern. For each feature, maintenance importance measures how strongly that feature distinguishes the patient’s assigned state from alternative states, calculated as the ratio of the distance to the assigned state centroid to the average distance to other state centroids. Features with high maintenance importance indicate strong specificity for the current state assignment.

For patients transitioning between states, transition importance measures how each feature contributed to movement from the previous state toward the new state. Transition importance combines changes in the feature that increase distance from the previous state centroid with simultaneous movement toward the new state centroid. Features exhibiting large, coordinated changes aligned with the transition direction receive higher scores. All feature importance scores were normalized to sum to 1.0 for each patient at each time snapshot, enabling interpretation as percentage contributions. This normalization ensured compatibility across patients with different state progression patterns and levels of clinical severity. Mathematical formulations are provided in Supplementary Methods S.M.3.

### Predictive Modeling and Performance Validation

#### Model Architecture and Feature Engineering

To evaluate the predictive capability of state-based features, we trained XGBoost gradient boosting models for mortality prediction across multiple prediction horizons (39). The feature matrix for each patient included 26 clinical variables (Table 1), 2 demographic variables (age, gender), and 6 state geometry descriptors (first state, last state, current Mahalanobis distance, and mean/minimum/maximum Mahalanobis distances), totaling 34 features.

#### Model training

Models employed binary logistic objective with conservative hyperparameters to limit overfitting: learning rate 0.05, maximum 64 leaves per tree, minimum child weight 6, L1 regularization (alpha) 0.8, and gamma 0.1. We used GPU-accelerated histogram-based tree construction for 2000 boosting rounds with early stopping patience of 100 rounds. We implemented patient-level train-test splits (80–20) with outcome stratification to ensure that no individual patient’s data appeared in both training and test sets. Class imbalance was addressed by setting the XGBoost scale_pos_weight parameter to (1−p)/p, where p represents the mortality rate in the training set. Complete hyperparameter details are provided in Supplementary Methods S.M.4.

#### Probability calibration

To ensure reliable probability estimates for clinical application (40), we reserved 20% of the training data as a calibration set and applied two post-hoc calibration methods: Platt scaling (logistic regression on model scores) and isotonic regression. Calibration quality was assessed using Expected Calibration Error (ECE), which measures the weighted average deviation between predicted probabilities and observed frequencies across probability bins (41), and Hosmer-Lemeshow goodness-of-fit test, which compares observed versus expected outcomes across risk deciles (42). Successful calibration was indicated by low ECE values (<0.05) and non-significant Hosmer-Lemeshow test results (p > 0.05).

#### Performance Metrics

Model discriminative performance was evaluated using area under the receiver operating characteristic curve (AUROC) and area under the precision-recall curve (AUPRC) on the held-out test set. We assessed performance for ICU mortality prediction across all five-time horizons using identical test patients.

### Cross Cohort Validation and Generalization

To assess generalizability, we applied the eICU-trained models to MIMIC-IV without retraining. Cross-dataset application required addressing systematic differences in measurement distributions. We implemented quantile-to-quantile distribution alignment (43). which maps each MIMIC-IV patient’s feature values to equivalent positions in the eICU-CRD reference distribution while preserving patient rankings. Following distribution alignment, MIMIC-IV snapshots were processed through the frozen state detection framework, with each snapshot assigned via Mahalanobis distance to eICU-derived state centroids. Hybrid feature importance scores were calculated identically to eICU-CRD processing.

We evaluated: (1) state-specific mortality rates compared with eICU-CRD patterns, (2) alignment-mortality relationships across ten alignment categories (0-10%, 10-20%, 20-30%, 30-40%, 40-50%, 50-60%, 60-70%, 70-80%, 80-90%, 90-100% time inside state boundaries), (3) predictive performance using AUROC and AUPRC, and (4) feature importance consistency using SHAP values. Detailed validation procedures, alignment quality metrics, and cross-dataset comparison analyses are provided in Supplementary Methods S.M.5.

## Implementation and Statistical Analysis

All analyses were executed in Python 3.12. Data manipulation was performed using Pandas for tabular operations and NumPy for vectorized numerical computations. Statistical functions were implemented using SciPy, including Wasserstein distance calculations for one-dimensional optimal transport, Mahalanobis distance for state assignment, cosine similarity for geometric comparisons, entropy calculations, chi-squared tests, and signal processing functions for peak detection during threshold optimization. The Python Optimal Transport (POT) library provided implementations of 2-Wasserstein distance for multidimensional population movement detection and Gromov-Wasserstein distance for internal geometric structure analysis within the 3-OT consensus voting system.

Scikit-learn provided preprocessing utilities, including median imputation for continuous variables and mode imputation for categorical variables, K-means clustering for threshold boundary detection, Gaussian Mixture Models for adaptive threshold estimation, and evaluation metrics including receiver operating characteristic curves, precision-recall curves, and calibration assessment tools. XGBoost gradient boosting models were trained using GPU-accelerated histogram-based tree construction. Feature importance was assessed using Shapely Additive exPlanations (SHAP) values calculated through TreeExplainer for gradient boosted models. Permutation-based feature importance with bootstrap resampling provided complementary importance estimates and confidence intervals.

Quantile-to-quantile transformation enabled distribution alignment between MIMIC-IV and eICU-CRD datasets while preserving patient rankings within each cohort. Patient-level train-test splits with outcome stratification ensured that no individual patient appeared in both training and evaluation sets, preventing data leakage. Bootstrap resampling (1000 iterations) was used to calculate 95% confidence intervals for performance metrics. Publication-quality figures were generated using Matplotlib and Seaborn with consistent styling conforming to journal specifications.

## Data Availability

The data used in this study are available from publicly accessible databases. The eICU Collaborative Research Database (eICU-CRD) version 2.0 is available at https://physionet.org/content/eicu-crd/2.0/. The Medical Information Mart for Intensive Care IV (MIMIC-IV) database is available at https://physionet.org/content/mimiciv/2.2/. Access to both databases requires completion of a credentialing process through PhysioNet, including human subjects research training and a signed data use agreement. The derived data supporting the findings of this study, including state assignments and model outputs, are available from the corresponding author upon reasonable request.

## SUPPLEMENTARY FIGURE LEGENDS

**Supplementary Figure 1. Calibration Analysis for the STREAM ICU Mortality Prediction Model.** (A) Reliability diagram comparing predicted probabilities with observed mortality for the original model (red), Platt scaling (green), and isotonic regression (blue); the dashed line indicates perfect calibration. (B) Predicted probability distributions showing concentration of the original model in very low-probability ranges (<0.1) and more appropriate spread after calibration. (C) Expected Calibration Error (ECE) comparison demonstrating substantial reduction in calibration error for both calibrated models relative to the original, confirming that post-hoc calibration successfully corrected probability estimates while maintaining identical discriminative performance (AUROC).

**Supplementary Figure 2. SHAP Feature Importance Analysis Across All Temporal Horizons for eICU-CRD Mortality Prediction.** (A) 8-hour model, (B) 24-hour model, (C) 48-hour model, (D) 72-hour model, and (E) Full ICU stay model. Violin plots display SHAP value distributions for the top 15 features at each prediction horizon, where values to the right increase predicted mortality and values to the left decrease risk; color indicates underlying feature value (blue = low, pink = high).

**Supplementary Figure 3. Cross-Dataset SHAP Analysis: eICU-Trained Models Applied to MIMIC-IV Data.** (A) 8-hour model, (B) 24-hour model, (C) 48-hour model, (D) 72-hour model, and (E) Full ICU stay model. Violin plots show SHAP value distributions for frozen eICU-trained XGBoost models applied to MIMIC-IV without retraining, demonstrating feature importance stability across datasets.

