## Supplementary Material and Method for "STREAM: State Trajectory Representation & Evolution-Aware Monitoring"

**SUPPLEMENTARY METHODS**

**S.M.1 Detailed Missing Data Imputation Procedures**

ICU data contain substantial missingness due to selective laboratory ordering based on clinical need. We used Multiple Imputation by Chained Equations (MICE) to estimate missing values while preserving relationships between variables (1). For each feature type, we applied appropriate imputation methods: continuous laboratory values used predictive mean matching (draws from observed values similar to predicted values), categorical variables used mode-based selection, ordinal variables used proportional odds models, and binary indicators used logistic regression. All imputations incorporated all other available features as predictors. For time-varying measurements like Glasgow Coma Scale and vital signs, we added temporal procedures: if a patient had some measurements, we filled interior gaps by linear interpolation and extended the most recent value forward or the earliest value backward to maintain clinical continuity.

After imputation, we standardized all features to comparable scales. Continuous laboratory values and vital signs were scaled to [0,1] using minimum and maximum values observed in the eICU-CRD cohort. Ordinal features (e.g., categorical laboratory interpretations) were mapped to [0,1] according to their natural ordering. Binary features remained as 0 or 1. All standardization parameters (minimum, maximum, means) were calculated exclusively from eICU-CRD training data (2).

**S.M.2 Mathematical Formulas for 3-OT Consensus Voting**

STREAM detects when the population transitions between physiological states by monitoring changes at three different levels: individual laboratory values, overall patient condition, and patient similarity patterns. We call this the 3-OT (Triple Optimal Transport) system, which operates like three independent judges examining the same data from different perspectives (3).

The Three Detection Methods:

OT-1D (Individual Feature Changes): 1-Wasserstein distance was computed for each of the 28 clinical features separately between consecutive snapshots (4). This univariate optimal transport metric quantifies distributional shifts in individual biomarkers across the population. Clinically, this detects feature-specific transitions such as systematic upward shifts in lactate distributions or widening of mean arterial pressure distributions.

OT-2D (Overall Population Movement): 2-Wasserstein distance was computed to measure movement in the complete 28-dimensional physiological space between consecutive snapshots (5). This multivariate optimal transport metric quantifies the minimum work required to transform one snapshot's patient distribution into the next. Clinically, this captures coordinated changes across multiple organ systems simultaneously.

OT-Geometric (Patient Relationship Changes): Gromov-Wasserstein distance was computed to measure changes in the internal geometric structure between patient populations across consecutive snapshots (6). This metric quantifies reorganization of patient similarity patterns independent of specific feature values. Clinically, this detects when the fundamental relationships between patients change.

Consensus Voting System:

Each method generates distances between consecutive snapshots. To determine which time points represent true state transitions rather than random fluctuations, each method independently applies three unsupervised threshold detection algorithms: KneeLocator (elbow detection) (7), Gaussian Mixture Models (cluster intersection) (8), and K-means clustering (midpoint boundaries) (9). Each algorithm produces a yes/no vote for each snapshot. The three algorithm votes are aggregated to produce one final vote per detection method. The final transition decision combines the three OT method votes using confidence-weighted consensus. Each method's vote receives a weight based on its signal quality:

*ConsensusScore(t) = Σᵢ wᵢ × cᵢ × vᵢ(t) / Σᵢ wᵢ × cᵢ*

where vᵢ(t)∈{0,1} is the binary vote from method i at snapshot t, wᵢ is the base weight, and cᵢ is the confidence score calculated from each method's signal dynamic range and consistency across snapshots. We record a state transition when ConsensusScore(t)>0.5, meaning at least two of the three methods agree.

Implementation for eICU-CRD: For the multicenter eICU-CRD population, we used the 0.5 consensus threshold (requiring agreement from at least 2 of 3 methods), acknowledging the inherent heterogeneity across hospitals and patient populations. This threshold balances sensitivity to detect meaningful transitions against specificity to avoid false positives from isolated fluctuations in single methods.

**S.M.3 Feature Importance Mathematical Framework**

Following state assignment at each time snapshot, we computed snapshot-specific feature importance scores to quantify each clinical variable's contribution to individual patient state progressions. We developed a hybrid importance scoring system that adapts to different state progression patterns while maintaining interpretability (10).

Hybrid Importance Framework: The system distinguishes between two primary clinical scenarios, each requiring a distinct analytical approach:

For patients maintaining the same state across consecutive time snapshots, we calculated maintenance importance to identify which features characterize their stable physiological pattern. For each feature f, maintenance importance quantifies how strongly that feature distinguishes the patient's assigned state from alternative states:

*Importance_maintenance(f) = |d_assigned(f)| / (|d_assigned(f)| + d̄_other(f))*

where d_assigned represented the absolute distance between the patient's feature value and the assigned state centroid for feature f, and d̄_other represented the average distance to all non-assigned state centroids for that feature. Features with high maintenance importance indicated strong specificity for the current state assignment.

For patients transitioning between states, we calculated transition importance to measure how each feature drove movement from the previous state toward the new state. Transition importance for feature f was calculated as:

*Importance_transition(f) = (Δ_away(f) + Δ_toward(f)) / Σ_all_features(Δ_away + Δ_toward)*

where Δ_away measured the change in feature f moving the patient's value away from the previous state centroid, and Δ_toward measured simultaneous movement toward the new state centroid. Features showing large, coordinated changes aligned with the transition direction received higher scores.

Normalization and Interpretation: All feature importance scores were normalized to sum to 1.0 for each patient at each time snapshot (11), enabling interpretation as percentage contributions. This normalization ensured that importance scores remained comparable across patients with different state progression patterns and clinical severity levels.

Implementation: At the first ICU snapshot, all patients received maintenance importance scores based on their initial state assignment. For subsequent snapshots, the system automatically determined whether to calculate maintenance importance (same state as previous snapshot) or transition importance (different state from previous snapshot). This hybrid approach ensured that every patient snapshot received an interpretable importance profile regardless of state progression stability or complexity.

**S.M.4 Complete XGBoost Hyperparameter Configuration and Training Details**

Model Architecture and Feature Engineering: To evaluate the predictive capability of state-based features, we trained XGBoost gradient boosting models for mortality prediction across multiple prediction horizons (12). The feature matrix for each patient included 26 clinical variables (Table 1), 2 demographic variables (age, gender), and 6 state geometry features (first state, last state, Mahalanobis distance, mean Mahalanobis distance, maximum Mahalanobis distance, minimum Mahalanobis distance), totaling 34 features.

We constructed prediction models at five time horizons: 8 hours (first snapshots only), 24 hours (pre-admission through first 24 hours), 48 hours (through 48 hours), 72 hours (through 72 hours), and full ICU stay (complete state progression data). For time-restricted models, all state-based features and state geometry metrics were computed exclusively from data within their respective time windows to prevent information leakage. Feature composition for each prediction horizon is detailed in Supplementary Table 2.

Model Training and Hyperparameters: Models employed binary logistic objectives with conservative hyperparameters selected to limit overfitting: learning rate 0.05, maximum 64 leaves per tree, minimum child weight 6, L1 regularization (alpha) 0.8, and gamma 0.1. We used GPU-accelerated histogram-based tree construction for 2000 boosting rounds with early stopping patience of 100 rounds. Random seed was fixed at 42 for reproducibility.

Data Splitting: We implemented patient-level train-test splits (80-20) with outcome stratification to ensure no individual patient's data appeared in both training and test sets. Within the training set, an additional patient-level split (80-20) was performed for hyperparameter optimization and threshold selection.

Missing Data and Class Imbalance: Missing values were imputed using median values for continuous variables and mode values for categorical variables, with imputation parameters derived exclusively from training data. Class imbalance was addressed by setting the XGBoost scale_pos_weight parameter to (1−p)/p, where p represents the mortality rate in the training set.

Decision Threshold Optimization: Optimal classification thresholds were determined using Youden's J statistic (maximizing sensitivity + specificity − 1) from validation set predictions, then applied consistently across all time-restricted scenarios to ensure fair comparison.

Probability Calibration: To ensure reliable probability estimates for clinical application (13), we reserved 20% of the training data as a calibration set and applied two post-hoc calibration methods: Platt scaling (logistic regression on model scores) and isotonic regression (with out-of-bounds extrapolation) (14, 15). Calibration quality was assessed on the held-out test set using Expected Calibration Error (ECE) measuring weighted average deviation between predicted probabilities and observed frequencies across probability bins, Hosmer-Lemeshow goodness-of-fit test comparing observed versus expected outcomes across risk deciles (16) ,and overall probability quality metrics including Brier score and log loss. Successful calibration was indicated by low ECE values (<0.05) and non-significant Hosmer-Lemeshow test results (p > 0.05).

Performance Metrics: Model discriminative performance was evaluated using area under the receiver operating characteristic curve (AUROC) and area under the precision-recall curve (AUPRC) on the held-out test set. We assessed performance for ICU mortality predictions across all five time horizons using identical test patients.

Baseline Model Comparison: To quantify the contribution of state geometry features, we trained a baseline XGBoost model using only 28 raw clinical features (26 clinical variables + 2 demographics) without state-based geometry features. This baseline model used identical hyperparameters, training procedures, and evaluation metrics. Comparison between STREAM and baseline models is presented in Supplementary Table 3.

**S.M.5 Detailed Cross-Dataset Validation Procedures**

Quantile-to-Quantile Distribution Alignment

To enable direct application of eICU-trained models to MIMIC-IV data, we implemented quantile-to-quantile distribution alignment to address systematic differences in measurement distributions between datasets for each of the 28 shared clinical variables, we constructed percentile ladders from the eICU-CRD reference distribution (0-100th percentiles). Each MIMIC-IV patient's feature value was located at its corresponding percentile within the MIMIC-IV distribution, then reassigned to the equivalent percentile value from the eICU-CRD distribution. This monotonic transformation preserved each patient's relative ranking within MIMIC-IV while standardizing absolute values to the eICU-CRD scale.

We assessed alignment quality using two metrics: Kolmogorov-Smirnov (KS) similarity scores comparing distribution shapes, and Spearman rank correlations confirming preservation of patient orderings. Across all 28 shared variables, the mapped MIMIC-IV distribution closely matched eICU-CRD reference distributions, with absolute percentile differences below 1.9% and Kolmogorov-Smirnov similarity scores exceeding 0.95 for two-thirds of variables. Critically, patient ranking was preserved: Spearman correlations exceeded 0.99 for 24 of 26 variables, confirming that the mapping maintained clinically meaningful relationships while enabling cross-dataset analysis.

State Assignment and Trajectory Mapping

Following distribution alignment, MIMIC-IV snapshots were processed through the state detection framework with all parameters frozen to prevent information leakage. Each snapshot received state assignment via Mahalanobis distance to the five eICU-derived state centroids. The state centroids, inverse covariance matrices, and 95% confidence boundaries were all computed exclusively from eICU-CRD data and applied without modification to MIMIC-IV.

Hybrid feature importance scores were calculated using the transition and maintenance importance algorithms applied identically to eICU-CRD processing. For patients maintaining the same state, maintenance importance quantified which features distinguished their assigned state from alternatives. For patients transitioning between states, transition importance measured which features drove the movement. All importance scores were normalized to sum to 1.0 for each patient at each time snapshot.

Geometric alignment metrics were computed for each patient, including percentage of time inside assigned state boundaries and Mahalanobis distance statistics (mean, minimum, maximum, and variance across all snapshots).

State-Specific Mortality Analysis

To assess generalizability of discovered states, we calculated mortality rates for patients assigned to each of the five states in MIMIC-IV and compared these patterns with eICU-CRD state-specific mortality rates. For each state, we identified all unique patients who entered that state at any time during their ICU course and calculated the proportion who died in the ICU. We evaluated whether the rank ordering of state mortality risk was preserved across datasets and whether absolute mortality rates remained within reasonable ranges given differences in patient populations and institutional practices.

State-specific mortality rates in MIMIC-IV demonstrated patterns consistent with eICU-CRD findings. State 5 again showed the highest mortality (11.6% in MIMIC-IV vs 10.1% in eICU-CRD), followed by State 1 (10.5% vs 9.0%). States 2 and 4 remained the lowest-risk states in both datasets (3.5% and 4.1% in MIMIC-IV vs 4.5% and 4.1% in eICU-CRD). State 3 showed slightly higher mortality in MIMIC-IV (8.6%) compared to eICU-CRD (6.6%). The overall pattern of elevated risk in States 1 and 5, with lower risk in intermediate states, replicated across both cohorts, supporting the generalizability of these state-specific prognostic profiles.

Alignment-Mortality Relationship Analysis

To test whether state outliers demonstrate elevated mortality risk in independent populations, we stratified MIMIC-IV patients into ten alignment categories based on percentage of time inside assigned state boundaries: 0-10%, 10-20%, 20-30%, 30-40%, 40-50%, 50-60%, 60-70%, 70-80%, 80-90%, and 90-100%. For each category, we calculated hospital mortality rates and compared these patterns with eICU-CRD alignment-mortality relationships to assess reproducibility.

The core finding that state outliers face elevated mortality replicated strongly in MIMIC-IV. State outliers (0-10% time inside boundaries) had 41.1% hospital mortality versus 5.8% for patients who remained inside state boundaries (90-100% inside boundaries), representing a 7.1-fold increase in risk. The relationship between alignment and mortality showed a general decreasing trend, with intermediate categories showing 31.5% (10-20%), 28.6% (20-30%), 36.8% (30-40%), 32.5% (40-50%), 25.3% (50-60%), 26.7% (60-70%), 19.9% (70-80%), and 14.0% (80-90%) mortality. This gradient closely paralleled the eICU-CRD pattern, where 0-10% alignment patients had 45.7% mortality compared with 5.1% for 90-100% alignment (9-fold difference).

Predictive Model Application

The frozen XGBoost models trained on eICU-CRD were applied directly to distribution-aligned MIMIC-IV feature matrices without retraining or hyperparameter adjustment. We reconstructed identical feature panels for all five time horizons: 8 hours, 24 hours, 48 hours, 72 hours, and full ICU stay. Each feature matrix included the same 34 features used in eICU-CRD: 28 clinical variables, 2 demographics (age, gender), and 6 state geometry features.

Model discriminative performance was evaluated using area under the receiver operating characteristic curve (AUROC) and area under the precision-recall curve (AUPRC) for ICU mortality predictions. For each prediction horizon, we calculated 95% confidence intervals using bootstrap resampling (1000 iterations). Cross-dataset performance was compared with eICU-CRD internal validation results to quantify generalization performance.

For ICU mortality prediction, the eICU-trained STREAM model achieved AUROCs of 0.798 at 8 hours, 0.815 at 24 hours, 0.836 at 48 hours, 0.857 at 72 hours, and 0.899 for full stay in MIMIC-IV. Compared to internal eICU-CRD validation performance, MIMIC-IV AUROCs were lower by 6.5 percentage points (8 hours), 5.8 points (24 hours), 5.3 points (48 hours), 4.5 points (72 hours), and 3.2 points (full stay). This degree of performance preservation during cross-dataset application without retraining demonstrates robust generalizability of the state-based risk features.

Baseline Model External Validation

To assess whether state geometry features improve generalization, we also applied the frozen baseline model (28 features without state geometry) to MIMIC-IV. The baseline model achieved AUROCs of 0.740 at 8 hours, 0.736 at 24 hours, 0.770 at 48 hours, 0.768 at 72 hours, and 0.789 for full stay. Compared to the STREAM model, the baseline showed substantially lower external performance, with STREAM outperforming baseline by 5.8–11.0 percentage points across all time horizons on MIMIC-IV. Notably, the performance gap between STREAM and baseline was larger on external validation than on internal validation, suggesting that state-based features capture more generalizable physiological patterns (Supplementary Table 3).

Feature Importance Analysis

To assess whether predictive drivers maintain consistent importance across populations, we calculated SHAP (SHapley Additive exPlanations) values for all eICU-trained models applied to MIMIC-IV test data. SHAP values provide model-agnostic explanations by quantifying each feature's contribution to individual predictions. For tree-based models like XGBoost, TreeExplainer provides exact SHAP values efficiently.

Feature importance rankings were compared between eICU-CRD and MIMIC-IV across all five prediction time horizons (8 hours, 24 hours, 48 hours, 72 hours, and full stay). For each time horizon, we extracted the top 10 most important features based on mean absolute SHAP values and assessed overlap between datasets. We evaluated whether state-based features (mean Mahalanobis distance, minimum Mahalanobis distance, maximum Mahalanobis distance, state assignments) maintained top-tier importance in the external cohort.

The top 10 features showed substantial overlap between eICU-CRD and MIMIC-IV across all prediction horizons. Geometric alignment metrics (mean and minimum Mahalanobis distance) remained in the top 4 for all models in both datasets. Clinical risk factors including age, Glasgow Coma Scale measures (minimum and maximum values), and state characteristics consistently appeared among the most important predictors in both datasets. This consistency indicates that the core drivers of state-based mortality prediction generalize across ICU populations despite differences in absolute feature rankings. The preservation of geometric alignment features as top predictors in an entirely independent dataset provides strong evidence that these features capture fundamental physiological patterns rather than dataset-specific artifacts.

**SUPPLEMENTARY RESULTS**

**S.R.1 Model Calibration Analysis**

Probability calibration is essential for clinical decision support, where predicted probabilities must reflect true outcome frequencies.We evaluated calibration using Expected Calibration Error (ECE) and Hosmer-Lemeshow goodness-of-fit tests across all prediction horizons.

Before calibration, the XGBoost models showed moderate miscalibration typical of tree-based methods, with ECE values ranging from 0.035 to 0.040 across time horizons. Platt scaling (logistic regression on model outputs) reduced ECE to 0.004--0.006, while isotonic regression achieved ECE values of 0.002--0.003. Both calibration methods produced non-significant Hosmer-Lemeshow test results (p > 0.05) for all models, indicating adequate fit between predicted and observed probabilities.

The full-stay model showed the best calibration (ECE = 0.002 after isotonic regression), while the 8-hour model required more aggressive calibration due to higher baseline miscalibration. Reliability diagrams demonstrated that calibrated probabilities fell within confidence bands across all probability ranges, with particular improvement in the high-risk (>0.5 predicted probability) regions most relevant for clinical alerting.

Brier scores decreased from 0.048–0.062 (uncalibrated) to 0.041–0.055 (calibrated), representing 8–15% improvement in overall probability quality. These results indicate that STREAM-based predictions can provide reliable probability estimates suitable for integration into clinical decision support systems.

**S.R.2 SHAP Feature Importance Analysis**

We analyzed feature importance using SHAP (SHapley Additive exPlanations) values to understand which features drive mortality predictions across different time horizons. SHAP values quantify each feature's contribution to individual predictions, enabling both global importance rankings and patient-level explanations.

Across all time horizons, state geometry features dominated the top importance rankings. Mean Mahalanobis distance (measuring average deviation from assigned state centroids) ranked as the most important feature for the 24-hour, 48-hour, 72-hour, and full-stay models, with SHAP importance values 2.1–3.4 times higher than the next most important feature. For the 8-hour model, minimum Mahalanobis distance ranked highest, reflecting the limited temporal data available at early time points.

Among clinical features, Glasgow Coma Scale minimum consistently appeared in the top 5 across all time horizons, confirming the prognostic importance of neurological status. Age ranked in the top 10 for all models. Lactate and creatinine showed increasing importance at longer time horizons, consistent with their roles as markers of organ dysfunction that accumulate over time.

The dominance of geometric alignment features (mean, minimum, and maximum Mahalanobis distance) suggests that the degree to which patients conform to expected physiological patterns provides stronger prognostic information than individual biomarker values alone. This finding supports the central thesis that state-based features capture clinically meaningful physiological patterns.

Feature importance remained stable across bootstrap resamples, with the top 5 features maintaining their rankings in >95% of iterations. This stability indicates that the importance hierarchy reflects genuine predictive relationships rather than sampling variability .

**S.R.3 Cross-Dataset Distribution Alignment Quality**

Successful cross-dataset validation requires that feature distributions are appropriately aligned between training (eICU-CRD) and external (MIMIC-IV) datasets. We evaluated alignment quality using multiple metrics.

Quantile-to-quantile transformation achieved excellent distribution matching. After transformation, Kolmogorov-Smirnov similarity scores exceeded 0.95 for 17 of 26 variables (65%) and exceeded 0.90 for 24 of 26 variables (92%). The two variables with lower similarity scores (lactate and anion gap) showed inherent measurement differences between datasets but maintained similarity scores above 0.85.

Patient ranking preservation was confirmed by Spearman rank correlations. For 24 of 26 variables, correlations between original MIMIC-IV rankings and transformed rankings exceeded 0.99, indicating that the transformation preserved relative patient orderings. The remaining two variables (GCS maximum and respiratory rate) showed correlations of 0.97 and 0.98 respectively, still demonstrating excellent preservation.

Clinical relationship preservation was assessed by examining correlations between related variables (e.g., creatinine-BUN, hemoglobin-hematocrit) before and after transformation. All clinically expected correlations were preserved with correlation coefficient changes <0.02, confirming that the transformation did not distort underlying physiological relationships.

**S.R.4 Detailed External Validation Results**

External validation in MIMIC-IV demonstrated robust generalization of STREAM-based mortality prediction. The frozen eICU-trained models achieved the following performance on MIMIC-IV without retraining:

8-hour model: AUROC 0.798 (95% CI: 0.796–0.799), representing a 6.5 percentage point decrease from eICU-CRD performance (0.863). Despite the larger performance drop at this early time point, the model still achieved clinically useful discrimination.

24-hour model: AUROC 0.815 (95% CI: 0.813–0.816), representing a 5.8 percentage point decrease from eICU-CRD (0.873).

48-hour model: AUROC 0.836 (95% CI: 0.835–0.838), representing a 5.3 percentage point decrease from eICU-CRD (0.890).

72-hour model: AUROC 0.857 (95% CI: 0.857–0.858), representing a 4.5 percentage point decrease from eICU-CRD (0.903).

Full-stay model: AUROC 0.899 (95% CI: 0.899–0.900), representing only a 3.2 percentage point decrease from eICU-CRD (0.932). This minimal performance drop demonstrates excellent generalization for the complete state-based model.

State-specific mortality patterns replicated across datasets. The rank ordering of state mortality risk was perfectly preserved: States 1 and 5 showed highest mortality in both datasets, while States 2 and 4 showed lowest mortality. Absolute mortality rates differed by <2 percentage points for 4 of 5 states.

State outlier mortality rates were elevated across all five states in eICU-CRD, regardless of whether the state itself was low-risk or high-risk: State 1 (23.9%), State 2 (56.7%), State 3 (23.2%), State 4 (31.0%), and State 5 (23.5%). The notably higher mortality rate observed for State 2 outliers (56.7%) likely reflects the small sample size of state outliers in this low-risk state (n=64), making this estimate susceptible to statistical variability rather than representing a true biological difference. This pattern replicated in MIMIC-IV with more consistent rates across states: State 1 (23.8%), State 2 (25.0%), State 3 (22.4%), State 4 (26.3%), and State 5 (22.6%), where larger sample sizes yielded more stable estimates. The consistency of state outlier mortality rates in MIMIC-IV (22-26% across all states) supports the finding that state outlier status confers elevated mortality risk independent of which state the patient is assigned to. The alignment-mortality relationship also replicated strongly. In MIMIC-IV, patients with 0-10% state alignment had 41.1% mortality versus 5.8% for patients with 90-100% alignment (7.1-fold difference). This closely paralleled the eICU-CRD pattern (45.7% vs 5.1%, 9-fold difference), confirming that state outliers face elevated mortality risk across independent populations.

**S.R.5 Cross-Dataset Feature Importance Comparison**

To assess whether predictive drivers generalize across datasets, we compared SHAP-based feature importance rankings between eICU-CRD and MIMIC-IV.

The top 5 features showed substantial overlap between datasets across all time horizons. Mean Mahalanobis distance appeared in the top 3 for both datasets across all models except 8-hour. Minimum Mahalanobis distance ranked in the top 5 for all models in both datasets. Age and Glasgow Coma Scale minimum consistently appeared in the top 10 for both datasets.

Correlation between feature importance rankings (using mean absolute SHAP values) was calculated for each time horizon. Spearman correlations ranged from 0.72 (8-hour) to 0.89 (full-stay), indicating strong consistency in which features drive predictions across datasets. The lower correlation at 8 hours reflects the limited temporal information available at early time points.

Notably, the three state geometry features (mean, minimum, and maximum Mahalanobis distance) maintained top-tier importance in MIMIC-IV despite being derived from state centroids learned exclusively on eICU-CRD. This provides strong evidence that these geometric features capture fundamental physiological patterns that generalize across institutions and patient populations.

**S.R.6 Baseline Model Comparison Results**

To quantify the contribution of state-based geometry features, we compared STREAM (34 features) with a baseline model using only raw clinical features (28 features: 26 clinical variables + 2 demographics) without state geometry features.

Internal Validation (eICU-CRD): STREAM consistently outperformed baseline across all time horizons. The improvement ranged from +5.5 percentage points (72 hours and full stay) to +6.7 percentage points (8 hours). At 8 hours, STREAM achieved AUROC 0.863 versus baseline 0.796. At full stay, STREAM achieved AUROC 0.932 versus baseline 0.877.

External Validation (MIMIC-IV): The performance advantage of STREAM over baseline was even larger on external validation. STREAM outperformed baseline by +5.8 percentage points at 8 hours (0.798 vs 0.740), +7.9 points at 24 hours (0.815 vs 0.736), +6.6 points at 48 hours (0.836 vs 0.770), +8.9 points at 72 hours (0.857 vs 0.768), and +11.0 points at full stay (0.899 vs 0.789).

Cross-Dataset Generalization: Importantly, STREAM showed better generalization than baseline. STREAM's cross-dataset performance decrease ranged from 3.2% (full stay) to 6.5% (8 hours), while baseline's decrease ranged from 5.6% (8 hours) to 8.9% (full stay). The baseline model's larger performance drop, particularly at longer time horizons, suggests that raw clinical features without state-based context are more susceptible to dataset-specific patterns.

These results demonstrate that state geometry features not only improve predictive performance but also enhance generalizability across institutions. The state-based features appear to capture more robust physiological patterns that transfer better to new populations than raw clinical measurements alone.

16.	Hosmer Jr DW, Lemeshow S, Sturdivant RX. Applied logistic regression: John Wiley & Sons; 2013.
