## Supplementary figures and images for "STREAM: State Trajectory Representation & Evolution-Aware Monitoring"

### Supplementary Figure 1.png

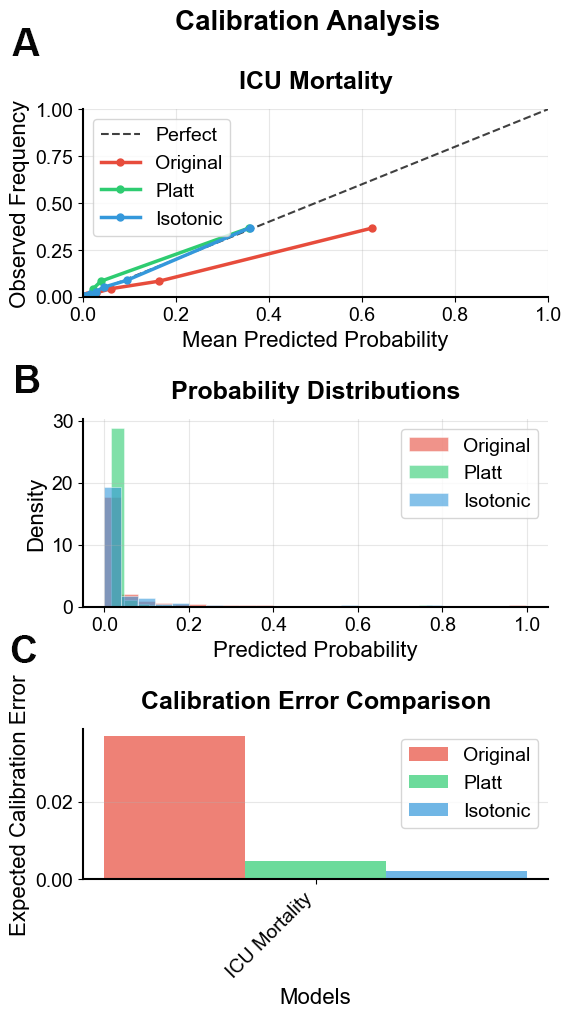

### Supplementary Figure 2.png

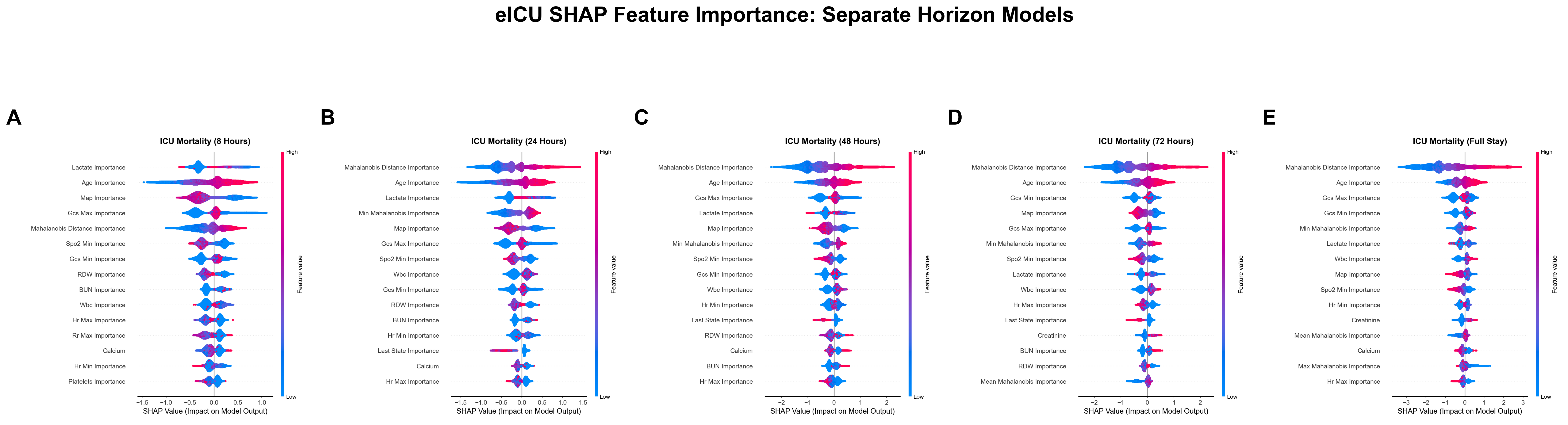

### Supplementary Figure 3.png

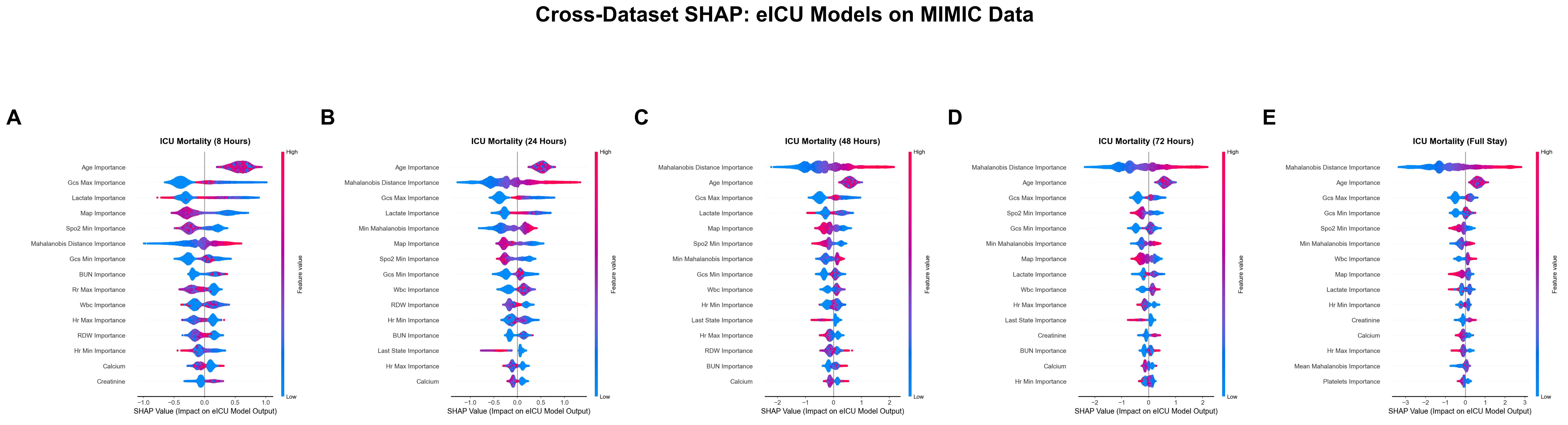
