## Supplementary Table 1 for "STREAM: State Trajectory Representation & Evolution-Aware Monitoring"

**Supplementary Table 1.** Cohort characteristics of eICU-CRD and MIMIC-IV datasets

| **Characteristic** | **eICU-CRD (N=158,294)** | **MIMIC-IV (N=84,517)** |
| --- | --- | --- |
| **Demographics** |  |  |
| Age, years, mean (SD) | 63.2 (16.4) | 65.1 (17.2) |
| Male sex, n (%) | 87,062 (55.0%) | 48,176 (57.0%) |
| **ICU Characteristics** |  |  |
| ICU length of stay, days, median (IQR) | 1.9 (1.0–3.8) | 2.1 (1.2–4.2) |
| Hospital length of stay, days, median (IQR) | 5.8 (3.2–10.4) | 6.4 (3.5–11.8) |
| **ICU Type, n (%)** |  |  |
| Medical ICU | 62,518 (39.5%) | 31,354 (37.1%) |
| Surgical ICU | 41,157 (26.0%) | 24,267 (28.7%) |
| Cardiac ICU | 33,242 (21.0%) | 16,903 (20.0%) |
| Neurological ICU | 12,660 (8.0%) | 5,918 (7.0%) |
| Other/Mixed ICU | 8,717 (5.5%) | 6,075 (7.2%) |
| **Outcomes, n (%)** |  |  |
| ICU mortality | 8,546 (5.4%) | 4,564 (5.4%) |
| Hospital mortality | 12,664 (8.0%) | 7,350 (8.7%) |
| **Severity Indicators, n (%)** |  |  |
| Mechanical ventilation | 47,488 (30.0%) | 28,975 (34.3%) |
| Vasopressor use | 31,659 (20.0%) | 19,398 (22.9%) |

*Abbreviations: SD, standard deviation; IQR, interquartile range; ICU, intensive care unit. Both cohorts included adult patients (≥18 years) with ICU stays exceeding 8 hours.*
