## Supplementary Table 2 for "STREAM: State Trajectory Representation & Evolution-Aware Monitoring"

**Supplementary Table 2.** Feature composition and data availability by prediction time horizon

| **Feature Category** | **8 hours** | **24 hours** | **48 hours** | **72 hours** | **Full Stay** |
| --- | --- | --- | --- | --- | --- |
| **Clinical Variables** | | | | | |
| Laboratory values (19 features) | ✓ | ✓ | ✓ | ✓ | ✓ |
| Vital signs (5 features) | ✓ | ✓ | ✓ | ✓ | ✓ |
| Neurologic status (2 features) | ✓ | ✓ | ✓ | ✓ | ✓ |
| **Demographics** | | | | | |
| Age | ✓ | ✓ | ✓ | ✓ | ✓ |
| Gender | ✓ | ✓ | ✓ | ✓ | ✓ |
| **State Geometry Features** | | | | | |
| First state assignment | ✓ | ✓ | ✓ | ✓ | ✓ |
| Last state assignment | – | ✓ | ✓ | ✓ | ✓ |
| Mahalanobis distance | ✓ | ✓ | ✓ | ✓ | ✓ |
| Mean Mahalanobis distance | – | ✓ | ✓ | ✓ | ✓ |
| Maximum Mahalanobis distance | – | ✓ | ✓ | ✓ | ✓ |
| Minimum Mahalanobis distance | – | ✓ | ✓ | ✓ | ✓ |
| **Data Availability** | | | | | |
| Snapshots included | 1 | 1–4 | 1–5 | 1–6 | 1–13 |
| Time coverage | 0–8h | 0–24h | 0–48h | 0–72h | 0–Day 10 |
| Total features | 31 | 34 | 34 | 34 | 34 |

*Check mark (✓) indicates feature is available at that time horizon; dash (–) indicates feature not available. All state geometry metrics were computed exclusively from data within the respective time window to prevent information leakage.*
