## Supplementary Table 3 for "STREAM: State Trajectory Representation & Evolution-Aware Monitoring"

**Supplementary Table 3.** Baseline model comparison: STREAM vs. raw clinical features

| **Time Window** | **Model** | **eICU-CRD AUROC (95% CI)** | **MIMIC-IV AUROC (95% CI)** | **Δ AUROC** |
| --- | --- | --- | --- | --- |
| 8 hours | Baseline | 0.796 (0.793–0.799) | 0.740 (0.738–0.742) | −0.056 |
|  | **STREAM** | **0.863 (0.860–0.866)** | **0.798 (0.796–0.799)** | **−0.065** |
|  | Δ (STREAM − Baseline) | +0.067 (+8.4%) | +0.058 (+7.8%) |  |
| 24 hours | Baseline | 0.810 (0.807–0.813) | 0.736 (0.734–0.738) | −0.074 |
|  | **STREAM** | **0.873 (0.868–0.878)** | **0.815 (0.813–0.816)** | **−0.058** |
|  | Δ (STREAM − Baseline) | +0.063 (+7.8%) | +0.079 (+10.7%) |  |
| 48 hours | Baseline | 0.833 (0.830–0.836) | 0.770 (0.768–0.772) | −0.063 |
|  | **STREAM** | **0.890 (0.886–0.893)** | **0.836 (0.835–0.838)** | **−0.053** |
|  | Δ (STREAM − Baseline) | +0.057 (+6.8%) | +0.066 (+8.6%) |  |
| 72 hours | Baseline | 0.848 (0.845–0.851) | 0.768 (0.766–0.770) | −0.080 |
|  | **STREAM** | **0.903 (0.898–0.908)** | **0.857 (0.857–0.858)** | **−0.045** |
|  | Δ (STREAM − Baseline) | +0.055 (+6.5%) | +0.089 (+11.6%) |  |
| Full Stay | Baseline | 0.877 (0.874–0.880) | 0.789 (0.787–0.791) | −0.088 |
|  | **STREAM** | **0.932 (0.930–0.933)** | **0.899 (0.899–0.900)** | **−0.032** |
|  | Δ (STREAM − Baseline) | +0.055 (+6.3%) | +0.110 (+13.9%) |  |

*Comparison of STREAM (34 features: 26 clinical + 2 demographics + 6 state geometry) with baseline model (28 features: 26 clinical + 2 demographics, no state geometry). Both models used identical XGBoost hyperparameters and training procedures. STREAM rows highlighted in green; improvement rows highlighted in yellow. Δ AUROC indicates cross-dataset performance change. STREAM showed larger improvement on external validation than internal validation, suggesting state-based features capture more generalizable patterns.*
