## Supplementary Table 4 for "STREAM: State Trajectory Representation & Evolution-Aware Monitoring"

**Supplementary Table 4. Cross-Dataset Distribution Alignment Quality**

| **A. Distribution Alignment Error Summary** | | |
| --- | --- | --- |
| **Metric** | **Value (%)** | **Feature** |
| Mean error | 4.9 | - |
| Median error | 2.8 | - |
| Minimum error | 0.8 | map_mean |
| Maximum error | 19.6 | gcs_max |
| **Feature-Level Error Distribution** | | |
| **Category** | **Feature** | **Error (%)** |
| Lowest-error | map_mean | 0.8 |
|  | wbc_x_1000 | 0.8 |
|  | platelets_x_1000 | 1.0 |
| Highest-error | gcs_max | 19.6 |
|  | gcs_min | 17.6 |
|  | spo2_min | 10.5 |

| **B. Clinical Relationship Preservation** | | | | |
| --- | --- | --- | --- | --- |
| **Relationship** | **Original** | **Rescaled** | **Preserved (%)** | **Quality** |
| Kidney function | 0.6247 | 0.6255 | 100.1 | Excellent |
| Anemia markers | 0.9654 | 0.9647 | 99.9 | Excellent |
| Electrolyte balance | -0.1808 | -0.1813 | 100.3 | Excellent |
| Heart rate variability | 0.6299 | 0.6342 | 100.7 | Excellent |
| Hemodynamic status | 0.0830 | 0.0701 | 84.4 | Fair |

| **C. Patient Ranking Preservation** | | | | |
| --- | --- | --- | --- | --- |
| **Feature** | **Spearman ρ** | **Pearson r** | **n** | **Quality** |
| calcium | 0.9995 | 0.9922 | 10,000 | Excellent |
| platelets_x_1000 | 1.0000 | 0.9994 | 10,000 | Excellent |
| chloride | 0.9994 | 0.9986 | 10,000 | Excellent |
| mch | 1.0000 | 0.9959 | 10,000 | Excellent |
| mcv | 1.0000 | 0.9988 | 10,000 | Excellent |

**Table caption:** Cross-dataset distribution alignment quality metrics for quantile-to-quantile transformation from MIMIC-IV to eICU-CRD. **(A)** Distribution alignment error summary showing mean, median, minimum, and maximum errors across all 28 features, with feature-level breakdown for lowest and highest error features. **(B)** Clinical relationship preservation showing correlation coefficients between clinically related features before (Original) and after (Rescaled) transformation. Preserved (%) indicates the proportion of original correlation maintained. **(C)** Patient ranking preservation showing Spearman and Pearson correlations between original and transformed values, confirming that patient orderings were maintained. Quality ratings: Excellent (≥95%), Good (85-94%), Fair (75-84%).

**Abbreviations:** map_mean = mean arterial pressure; gcs_max/min = Glasgow Coma Scale maximum/minimum; wbc = white blood cell count; spo2_min = oxygen saturation minimum; mch = mean corpuscular hemoglobin; mcv = mean corpuscular volume.
